# Mask Wearing and Control of SARS-CoV-2 Transmission in the United States

**DOI:** 10.1101/2020.08.23.20078964

**Authors:** Benjamin Rader, Laura F. White, Michael R. Burns, Jack Chen, Joe Brilliant, Jon Cohen, Jeffrey Shaman, Larry Brilliant, Moritz U.G. Kraemer, Jared B. Hawkins, Samuel V. Scarpino, Christina M. Astley, John S. Brownstein

## Abstract

**Introduction:** Cloth face coverings and surgical masks have become commonplace across the United States in response to the SARS-CoV-2 epidemic. While evidence suggests masks help curb the spread of respiratory pathogens, population level, empirical research remains is limited. Face masks have quickly become a topic of public debate as government mandates have started requiring their use. Here we investigate the association between self-reported mask wearing, social distancing and community SARS-CoV-2 transmission in the United States, as well as the effect of statewide mandates on mask uptake.

**Methods:** Serial cross-sectional surveys were administered June 3 through July 27, 2020 via web platform. Surveys queried individuals’ likelihood to wear a face mask to the grocery store or with family and friends. Responses (N=378,207) were aggregated by week and state and combined with measures of the instantaneous reproductive number (*R_t_*), social distancing proxies, respondent demographics and other potential sources of confounding. We fit multivariate logistic regression models to estimate the association between mask wearing and community transmission control (*R_t_* <1) for each state and week. Multiple sensitivity analyses were considered to corroborate findings across mask wearing definitions, *R_t_* estimators and data sources. Additionally, mask wearing in 12 states was evaluated two weeks before and after statewide mandates.

**Results:** We find an increasing trend in mask usage across the U.S., although uptake varies by geography and demographic groups. A multivariate logistic model controlling for social distancing and other variables found a 10% increase in mask wearing was associated with a 3.53 (95% CI: 2.03, 6.43) odds of transmission control (*R_t_* <1). We also find that communities with high mask wearing and social distancing have the highest predicted probability of a controlled epidemic. These positive associations were maintained across sensitivity analyses. Following state mandates, mask wearing did not show significant statistical changes in uptake, however the positive trend of increased mask wearing over time was preserved.

**Conclusion:** Widespread utilization of face masks combined with social distancing increases the odds of SARS-CoV-2 transmission control. Mask wearing rose separately from government mask mandates, suggesting supplemental public health interventions are needed to maximize mask adoption and disrupt the spread of SARS-CoV-2, especially as social distancing measures are relaxed.

## Introduction

In December 2019, SARS-CoV-2 and the resulting COVID-19 disease were first identified in Wuhan, China^1^. The disease has spread globally since its identification, causing widespread morbidity and mortality^2,3^. In the absence of a vaccine or curative therapy, governments in the United States and worldwide have adopted numerous approaches to curb the virus’s continued transmission^4,5^. Despite widespread implementation, the efficacy of various nonpharmaceutical interventions has been intensely debated^6^, resulting in significant individual and community heterogeneity in the acceptance of these interventions at the individual and community level ^7^, including the use of face masks and respirators^8^.

Evidence strongly supports N95 respirators as an effective method to prevent viral respiratory transmission, but supply chain shortages necessitate their preferential allocation to high-risk, front-line medical personnel^9,10^. Consequently, cloth face coverings and surgical masks (henceforth collectively, “face masks”) are recommended as an alternative for the general public^11^. Following the initial spread of the virus in the United States, many local and state jurisdictions have mandated the use of face masks in public settings^5^. These masks are intended to serve as a mechanical barrier that absorbs virus-laden droplets expelled by the user^12^. Therefore, their purpose is to reduce transmission events by the user, rather than protect the individual from infection. Accordingly, face masks are advocated as a source of collective benefit, akin to herd immunity with vaccination, which is most successful with high levels of adoption^13^.

Empirical, population level evidence on the effectiveness of face masks at preventing respiratory transmission of SARS-CoV-2 is limited but growing^11,14^. While work on other respiratory infections^15^ and two recent case reports^16,17^ suggest mask usage may be effective, a recent global analysis found a limited marginal effect of mask mandates in the presence of other interventions^18^. Here, we use an ecological approach to assess mask compliance directly, irrespective of mandates. We combine survey data on personal mask wearing habits across the United States with a time varying measure of transmission control, as quantified by the effective reproductive number (*R_t_*) in each state. We then evaluate the association of a change in mask wearing with the timing of mask mandates to better understand their impact on transmission of COVID-19.

## Materials and Methods

### Survey Description

A web survey hosted on SurveyMonkey.com was deployed and promoted in conjunction with COVIDNearYou (a Boston Children’s Hospital based digital surveillance platform), as part of an effort to increase participatory syndromic surveillance for COVD-19^19–21^. The survey data were collected through SurveyMonkey’s “end page river sampling” [www.surveymonkey.com/mp/survey-methodology]. Briefly, over 2 million people around the world complete surveys designed by individuals, community groups, and businesses using the SurveyMonkey online survey platform every day. At the completion of these surveys, respondents in the United States are randomly offered to participate in the COVIDNearYou web survey. The river sample is not a stochastic probability sample of the entire population; however, it reaches respondents with diverse geographic and demographic backgrounds to ensure broad representativeness. Responses to this questionnaire were collected between June 3, 2020 and July 27, 2020 in all 50 states and the District of Columbia. Respondents were not provided incentives to complete the questionnaire. This study was approved by the Boston Children’s Hospital Institutional Review Board.

Survey responses were analyzed using crude data (unweighted) and survey weights that reflect the demographic composition of the United States. Survey weights standardize for age, race, sex, education, and geography using the Census Bureau’s American Community Survey. Also included was an additional smoothing parameter for political party identification based on aggregates of SurveyMonkey research surveys, refreshed weekly with rolling two-week aggregated data. Weights are generated daily for weekday surveys and once after weekend surveys.

### Mask Wearing Exposure

Survey respondents were asked a range of questions (full survey: www.surveymonkey.com/r/COVIDNearYouSurveyMonkey) including how likely they were to wear a mask while “grocery shopping” or “visiting family and friends” on a 4-point scale, from “very likely” to “not likely at all”. A composite exposure of consistent self-reported face mask wearing (referred to as *mask wearing*) is defined as the percentage of respondents who replied “very likely” to both questions. The percentage of respondents who replied “very likely” to each question, separately, was also evaluated in sensitivity analyses. Surveys without responses to either question (N=4,186) were excluded. For validation, the exposure was aggregated by county (limited to those with over 50 observations, N = 324) between July 2 and July 14, 2020 and compared against cross-sectional New York Times and Dynata interviews on mask wearing completed during this time period. ^8^

### Social Distancing Exposure

Population-level social distancing by state and by week was quantified as the duration of time spent at home compared to baseline (defined as January 3, 2020 - February 6, 2020)^22^. Duration of time spent at home was estimated with the Google community “residential time” mobility measure^23^, which was estimated using anonymized and aggregated data from individual Google users who opted into location history on their mobile devices^24^. A measure of social distancing from Facebook’s COVID-19 symptom survey^25^ was included in a sensitivity analysis. Individuals were asked “In the past 24 hours, with how many people have you had direct contact, outside of your household,” for each of a variety of settings. The number of self-reported contacts at “social gatherings” (censored outlier responses) was aggregated over each week and state utilizing Facebook’s weighted sampling scheme^25^.

### Community Transmission Control Outcome

The daily estimated instantaneous reproductive number (*R_t_*) – the number of secondary cases arising from a single case for a given day – was used to measure state-specific community transmission control. *R_t_* was aggregated to the week and dichotomized as epidemic slowing (1 if *R_t_* < 1) or maintenance/growth (0 if *R_t_* ≥ 1). *R_t_* estimates were extracted from rt.live, which was fit to case data from The COVID Tracking Project and the open COVID-19 data working group (method and adaptation previously described^3,26-28^). Sensitivity analyses were performed with *R_t_* values extracted from epiforecasts.io^29,30^ (available through July 19, 2020) and with *R_t_* dichotomized at different cutoffs.

### Modelling Mask Effectiveness on Transmission Control

We fit multivariate logistic regression models using *R* (version 3.6.2) to predict the community transmission control outcome (binary *R_t_*) using state- and week-specific estimates of mask wearing (crude and survey-weighted) and social distancing (relative residential time). State population density was included as a potential confounder given the association between population structure and SARS-CoV-2 transmission^31,32^ as well as the association between urban versus rural regions and face mask usage^8^. Percent non-white was included as a confounder due to the relationship with epidemiological indicators of SARS-CoV-2^33^ and uptake of non-pharmaceutical interventions^34^. A linear weekly time trend was also modeled.

To address the potential of reverse causation, where high previous transmission induced elevated mask wearing as well as lower potential *R_t_* (due to reduced effective contact availability), we included each state’s peak *R_t_* from March-May 2020 as a confounder in a sensitivity analysis. We also evaluated an interaction between the exposures of mask wearing and social distancing.

For each model, influential observations (up to n = 24) with a Cook’s distance over 4/N were excluded^35^. The influential observations removed varied between models, except for observations from New Jersey (characterized by high mask wearing, very high social distancing, and high community transmission control) which were repeatedly excluded for influence. Serial correlation was assessed in the final models but was abated following the aggregation of data.

Two additional modeling frameworks were utilized in sensitivity analyses. While each survey consisted of an independent sample of respondents, a mixed model with a random intercept for state and the same fixed effects as reported in the crude logistic model (model 1) was fit to account for the potential hierarchical structure of the observations. Additionally, an ordinal logistic regression was fit to measure the association of mask wearing with multiple, ordered categories of *R_t_*.

### Mask Mandates

The date for each statewide mask mandate was extracted from the masks4all.org database^36^. To assess the effect of mask mandates on mask wearing, segmented regression^37^ was run comparing the 2 weeks preceding and 2 weeks following each state’s intervention.

## Results

Self-reported mask wearing was evaluated using N=378,207 survey responses recorded between June 3, 2020 and July 27, 2020. Most (84.6%) reported “very likely” to wearing a face mask to the grocery store, while just under half (40.2%) did so to visit friends and family. A similar fraction (39.8%) reported they were very likely to wear a mask to the grocery store and with family and friends. Very few (4.7%) reported they were “not likely [to wear a mask] at all” in either setting. The percentage of individuals in each county who reported “always” wearing a mask in the New York Times interviews was correlated with those who reported they were “very likely” to wear a mask to the grocery store (Spearman’s *ρ* = .80, *p*<0.0001), with family and friends (Spearman’s *ρ* = .57, *p*<0.0001), and the composite score (Spearman’s *ρ* = .57, *p*<0.0001).

Mask wearing was higher among women, elderly, non-white or Hispanic, and lower income respondents (*p*<0.0001, **Table 1**). There was substantial geographic heterogeneity in survey responses (**Figure 1**, **A**.), with the highest percentage of mask wearers along the coasts and southern border, as well as in large urban areas. There was a general trend of increased mask use across U.S. Census Divisions throughout the survey period (**Figure 1**, **B**.) and the West North Central Census Division reported the lowest mask usage across the entire period surveyed.

**Table 1.** Characteristics of Unweighted Survey Respondents by Mask Wearing Status Characteristics of Unweighted Survey Respondents by response to the question, “how likely are you to wear a face mask to visit family and friends.” For each response, N (row %) and p-value from χ² test is presented. Overall N (column %) also included for each category.

|  | **Very Likely**<br>**No. 152,158** | **Somewhat Likely**<br>**No. 81,596** | **Not So Likely**<br>**No. 73,723** | **Not Likely At All**<br>**No. 67,832** | **Total**<br>**No. 375,309** | **P-value** |
| --- | --- | --- | --- | --- | --- | --- |
| Sex |  |  |  |  |  | < 0.0001 |
| Female | 103,624 (42%) | 54,314 (22%) | 47,825 (19%) | 40,908 (17%) | 246,671 (66%) |  |
| Male | 46,543 (38%) | 26,374 (21%) | 25,226 (20%) | 25,910 (21%) | 124,053 (33%) |  |
| Missing | 1,991 (43%) | 908 (20%) | 672 (15%) | 1,014 (22%) | 4,585 (1%) |  |
| Age |  |  |  |  |  | < 0.0001 |
| 13-17 | 7 (33%) | 4 (19%) | 5 (24%) | 5 (24%) | 21 (0%) |  |
| 18-24 | 6,328 (33%) | 4,626 (24%) | 4,564 (24%) | 3,895 (20%) | 19,413 (5%) |  |
| 25-34 | 15,943 (35%) | 9,790 (22%) | 9,955 (22%) | 9,629 (21%) | 45,317 (12%) |  |
| 35-44 | 25,679 (36%) | 14,938 (21%) | 14,339 (20%) | 15,503 (22%) | 70,459 (19%) |  |
| 45-54 | 32,138 (40%) | 17,505 (22%) | 15,778 (19%) | 15,755 (19%) | 81,176 (22%) |  |
| 55-64 | 35,005 (44%) | 17,697 (22%) | 14,915 (19%) | 12,772 (16%) | 80,389 (21%) |  |
| 65+ | 35,763 (48%) | 15,874 (21%) | 13,114 (18%) | 9,372 (13%) | 74,123 (20%) |  |
| Missing | 1,295 (29%) | 1,162 (26%) | 1,053 (24%) | 901 (20%) | 4,411 (1%) |  |
| Race/Ethnicity |  |  |  |  |  | < 0.0001 |
| White | 91,105 (35%) | 58,395 (22%) | 58,303 (22%) | 55,380 (21%) | 263,183 (70%) |  |
| Black | 26,904 (62%) | 8,638 (20%) | 4,579 (11%) | 2,955 (7%) | 43,076 (11%) |  |
| Hispanic | 8,858 (57%) | 3,237 (21%) | 2,166 (14%) | 1,309 (8%) | 15,570 (4%) |  |
| Other | 16,510 (49%) | 7,314 (22%) | 5,513 (16%) | 4,202 (13%) | 33,539 (9%) |  |
| Missing | 8,781 (44%) | 4,012 (20%) | 3,162 (16%) | 3,986 (20%) | 19,941 (5%) |  |
| Household Income |  |  |  |  |  | < 0.0001 |
| <\$50,000 | 47,847 (44%) | 23,025 (21%) | 19,373 (18%) | 17,927 (17%) | 108,172 (29%) | |
| \$50,000-\$99,000 | 43,380 (39%) | 24,601 (22%) | 22,285 (20%) | 20,707 (19%) | 110,973 (30%) | |
| >\$100,000 | 51,010 (38%) | 29,095 (22%) | 27,951 (21%) | 25,954 (19%) | 134,010 (36%) | |
| Missing | 9,921 (45%) | 4,875 (22%) | 4,114 (19%) | 3,244 (15%) | 22,154 (6%) |  |
| Census Division |  |  |  |  |  | < 0.0001 |
| New England | 10,747 (48%) | 5,421 (24%) | 3,838 (17%) | 2,518 (11%) | 22,524 (6%) |  |
| Middle Atlantic | 20,939 (45%) | 10,336 (22%) | 8,407 (18%) | 6,826 (15%) | 46,508 (12%) |  |
| East North Central | 17,206 (35%) | 11,205 (23%) | 10,723 (22%) | 10,117 (21%) | 49,251 (13%) |  |
| West North Central | 7,692 (28%) | 6,168 (23%) | 6,557 (24%) | 6,762 (25%) | 27,179 (7%) |  |
| East South Central | 7,508 (35%) | 4,484 (21%) | 4,530 (21%) | 4,838 (23%) | 21,360 (6%) |  |
| West South Central | 15,332 (40%) | 7,883 (21%) | 7,490 (20%) | 7,417 (19%) | 38,122 (10%) |  |
| South Atlantic | 34,688 (42%) | 17,453 (21%) | 15,610 (19%) | 14,218 (17%) | 81,969 (22%) |  |
| Pacific | 27,244 (48%) | 11,989 (21%) | 9,878 (17%) | 8,028 (14%) | 57,139 (15%) |  |
| Mountain | 10,802 (35%) | 6,657 (21%) | 6,690 (21%) | 7,108 (23%) | 31,257 (8%) |  |

**Figure 1.**
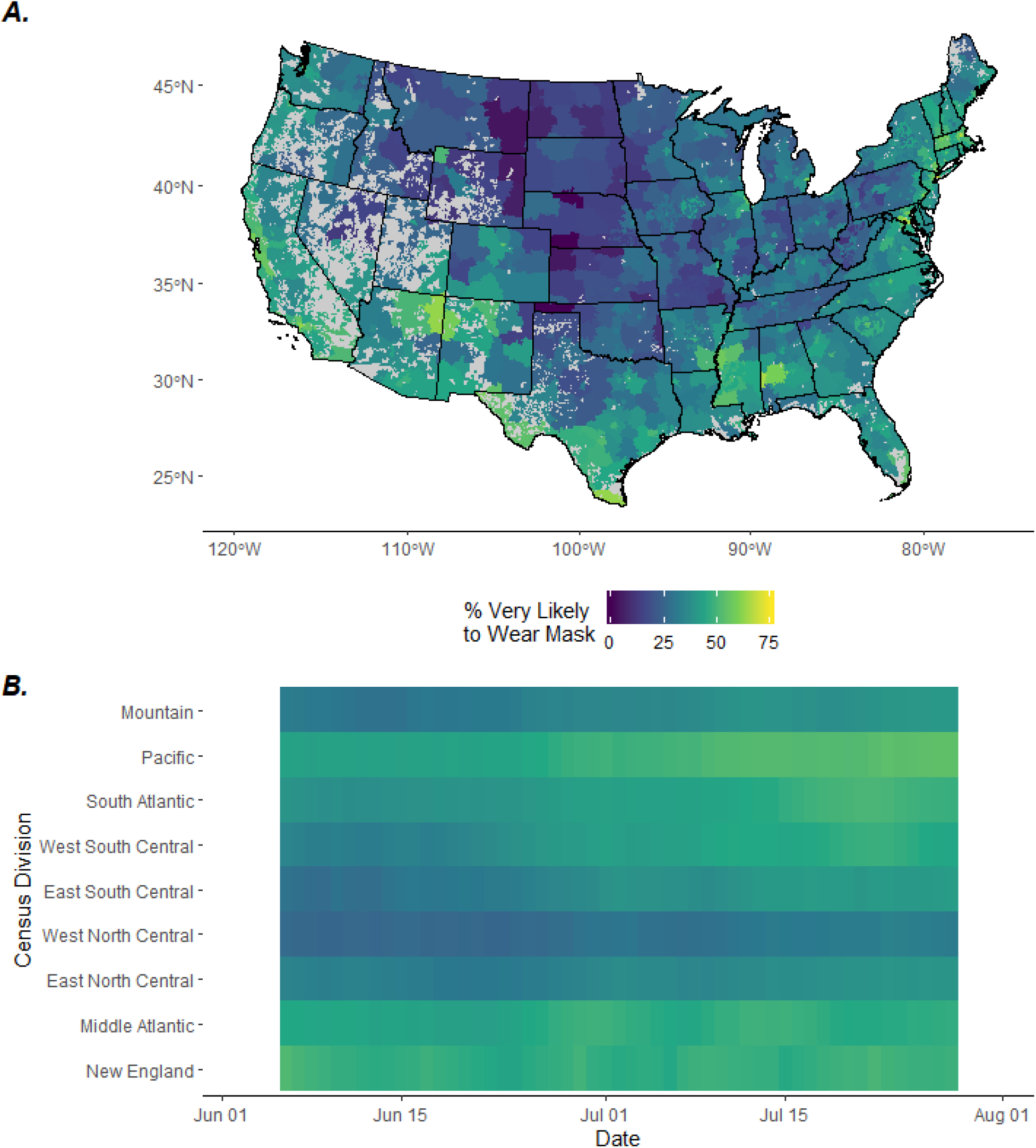
Mask wearing across three-digit zip code and time The percentage of individuals who responded they were very likely to wear a mask to the grocery store and with family and friends was averaged across the entire study period and each 3-digit ZCTA prefix (fuzzy matched with reported zip code) in the United States (Panel A). ZCTA clusters with less than 50 responses are shown in gray. Observations of mask wearing were also aggregated at the daily level across each Census division for the study period (Panel B).

We find a negative relationship between the mean percentage of people that report wearing a mask and the instantaneous reproductive number (**Figure 2)**. In a multivariate logistic regression model adjusting for confounders, social distancing and a time trend, there is a significant association between percent reporting mask wearing and community transmission control (i.e. *R_t_* <1) with an OR = 1.14 [95% CI: 1.07,1.20] (**Table 2, Model 1**). A 10% increase in mask wearing was associated with an over three-fold increase in odds of transmission control [OR = 3.53, 95% CI: 2.03, 6.43]. The association between mask wearing and community transmission control was robust to changes in definitions of mask wearing, *R_t_* estimated from epiforecasts.io, control for peak *R_t_*, and survey-weight standardization (**Table 2, Models 2-6**). When adjusting for community self-reported contacts instead of mobility, the association between one percent change in mask wearing and community transmission control was attenuated [OR = 1.09, 95% CI: 1.02, 1.16], but remained significant (**Figure S1**). Mask wearing was also significantly associated with reduced transmission across multiple *R_t_* dichotomization thresholds (**Figure S2**) and when categorized as an ordinal variable (**Figure S3**). A mixed model with a random intercept for state found a stronger association of mask wearing on community transmission control [OR = 1.18, 95% CI: 1.07, 1.30].

**Table 2.** Regression Results Estimated odds ratios [95% confidence intervals] from multivariable logistic regression models. Model 1 reports the association of the outcome of community transmission control (*R_t_* <1) with the percentage of surveys reporting “very likely” to wear a mask to the grocery store and to visit family and friends, aggregated by state and week. Model 1 controls for a weekly time trend, social distancing (relative Google mobility “residential time”), percentage non-white and population density. We evaluated the association using survey weights (Model 2), alternative estimators of *R_t_* (Model 3), and alternative definitions of mask usage (Models 4 and 5). Model 6 accounts for each states peak *R_t_* prior to the start of the study and model 7 includes an interaction of mask wearing with social distancing.

| | 1. Association with $R_t$ (rt.live) | 2. Weighted Sample | 3. Association with $R_t$ (epiforecasts) | 4. Alternative Mask Definition 1 | 5. Alternative Mask Definition 2 | 6. Accounting for Prior Transmission | 7. With Interaction Term |
| --- | --- | --- | --- | --- | --- | --- | --- |
| Mask Wearing (% , Composite) | 1.14 ***<br>[1.07, 1.20] | 1.13 ***<br>[1.07, 1.20] | 1.10 ***<br>[1.04, 1.15] |  |  | 1.14 ***<br>[1.08, 1.21] | 1.17 **<br>[1.05, 1.32] |
| Time Trend (Week) | 1.36 ***<br>[1.17, 1.58] | 1.35 ***<br>[1.16, 1.57] | 1.27 ***<br>[1.11, 1.45] | 1.37 ***<br>[1.18, 1.59] | 1.40 ***<br>[1.20, 1.63] | 1.38 ***<br>[1.18, 1.60] | 1.35 ***<br>[1.17, 1.56] |
| Social Distancing (% , Proxy) | 1.42 ***<br>[1.16, 1.74] | 1.43 ***<br>[1.17, 1.75] | 1.37 **<br>[1.13, 1.66] | 1.41 ***<br>[1.15, 1.72] | 1.52 ***<br>[1.24, 1.86] | 1.39 **<br>[1.13, 1.72] | 1.77<br>[0.99, 3.15] |
| Non-White (%) | 0.94 ***<br>[0.91, 0.96] | 0.94 ***<br>[0.91, 0.97] | 0.94 ***<br>[0.92, 0.97] | 0.94 ***<br>[0.91, 0.96] | 0.96 ***<br>[0.93, 0.98] | 0.93 ***<br>[0.90, 0.96] | 0.95 ***<br>[0.92, 0.97] |
| Density (1000 people/sq mi) | 0.09 **<br>[0.02, 0.39] | 0.08 ***<br>[0.02, 0.31] | 0.30 *<br>[0.10, 0.88] | 0.09 **<br>[0.02, 0.39] | 0.08 ***<br>[0.02, 0.32] | 0.03 ***<br>[0.01, 0.15] | 0.10 **<br>[0.03, 0.41] |
| Mask Wearing (% , With Friends) |  |  |  | 1.14 ***<br>[1.08, 1.21] |  |  |  |
| Mask Wearing (% , Grocery) |  |  |  |  | 1.07 **<br>[1.02, 1.12] |  |  |
| State Peak $R_t$ (March - May) | | | | | | 2.60 ***<br>[1.49, 4.56] | |
| Mask*Social Distancing |  |  |  |  |  |  | 0.99<br>[0.98, 1.01] |
| N | 376 | 379 | 378 | 376 | 382 | 381 | 382 |
| AIC | 347.78 | 352.83 | 383.84 | 347.10 | 366.59 | 346.94 | 363.63 |
| Pseudo R2 | 0.39 | 0.38 | 0.31 | 0.40 | 0.35 | 0.41 | 0.37 |

\*\*\* p < 0.001; \*\* p < 0.01; \* p < 0.05.

**Figure 2.**
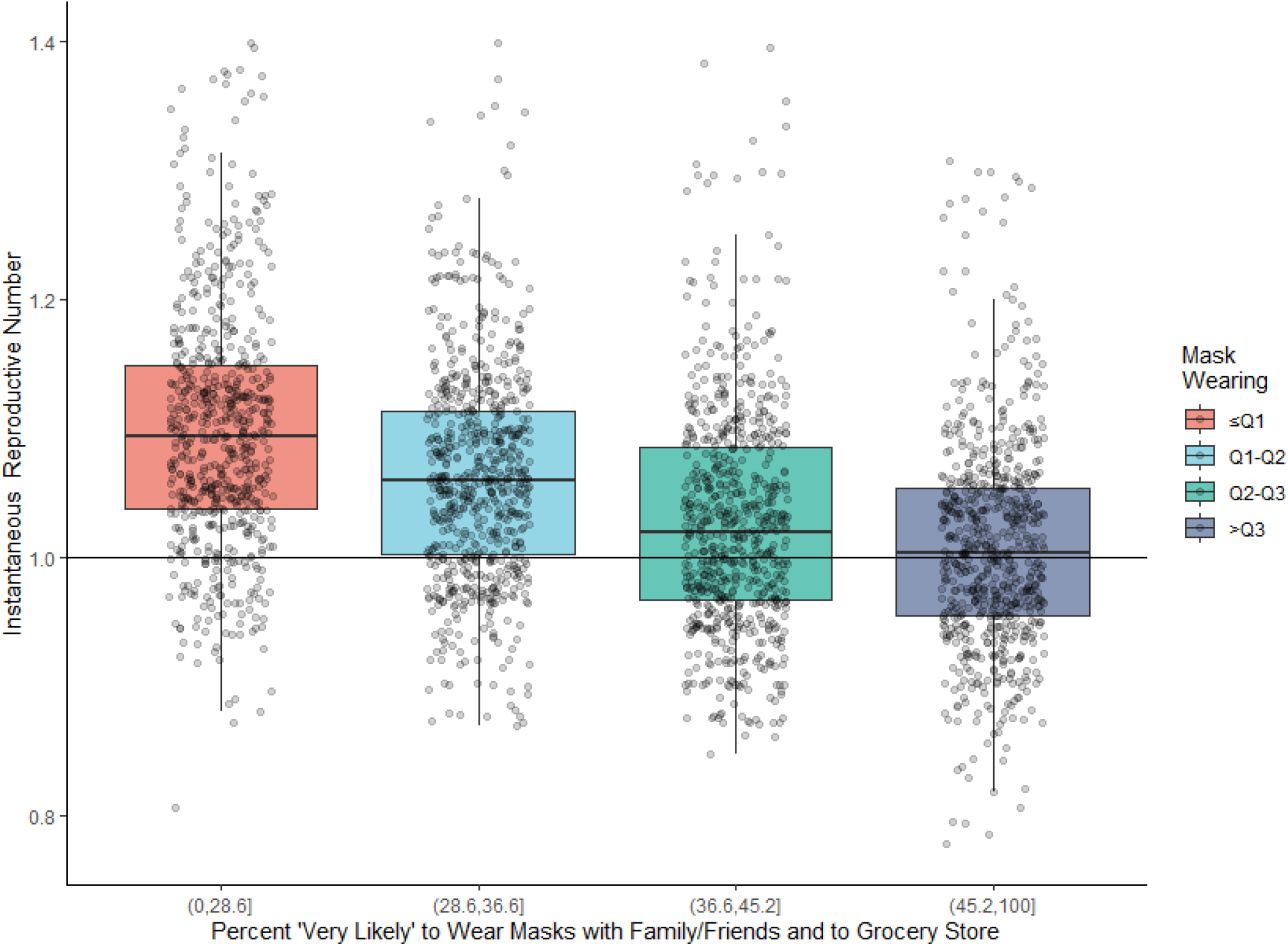
Mask Wearing and the Effective Reproductive Number Box (median and IQR) and whiskers (minimum and maximum excluding outliers) of *R_t_* estimates (rt.live) for each week and state (grey circles). Plot is stratified by quartiles (Q1: 28.8%, Q2: 36.6%, and Q3: 45.2%) of the percentage of individuals that report they are very likely to wear a mask with family/friends and to the grocery store.

Communities with high levels of mask wearing and social distancing are predicted by the model to have the highest probability of community transmission control (**Figure 3**). States with the highest percentage of reported mask wearing but lowest levels of social distancing have less than a 35% probability of community transmission control, though there are wide confidence intervals. The interaction between social distancing and mask wearing was not statistically significant (**Table 2, Model 7**).

**Figure 3.**
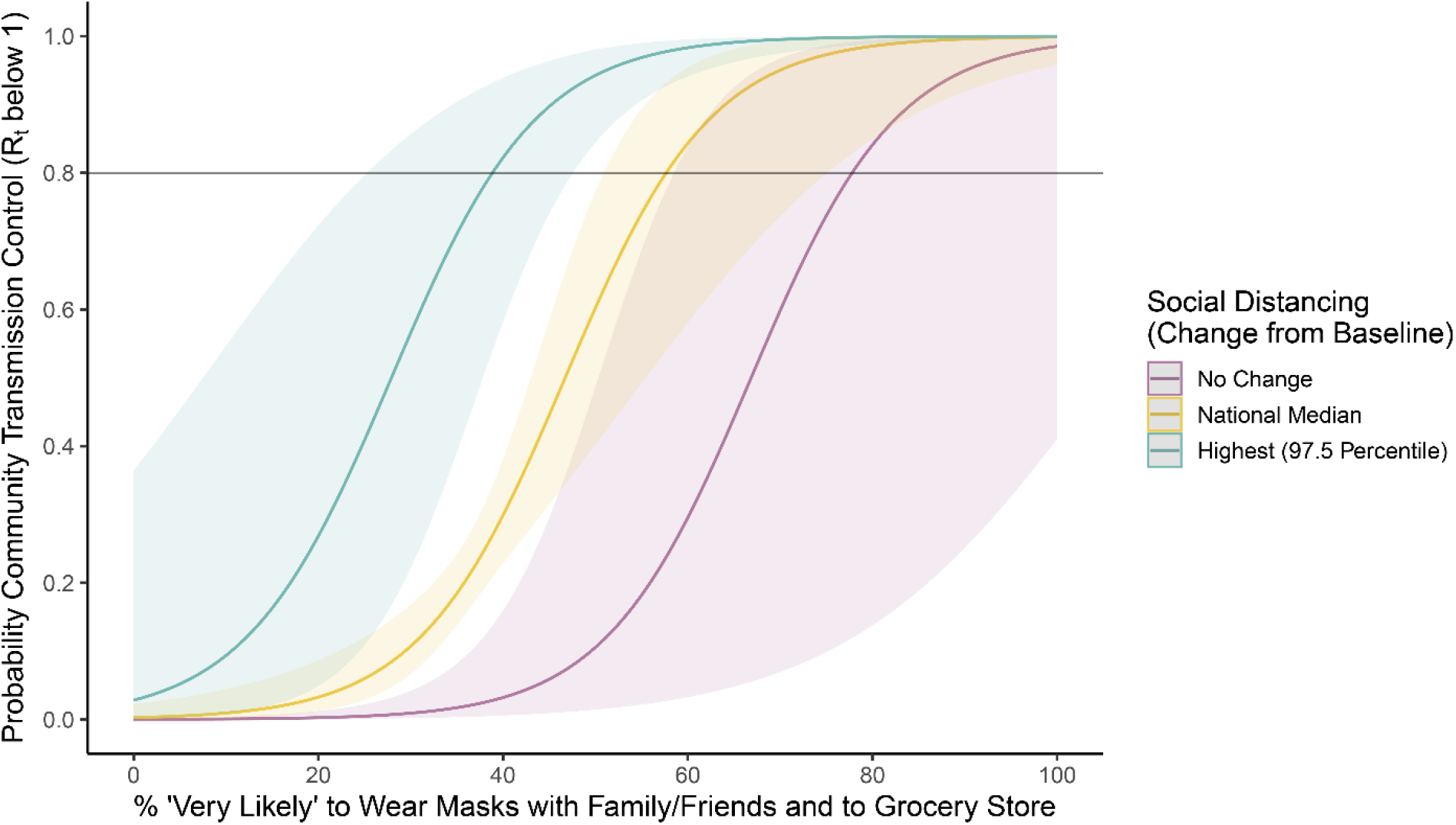
Mask Wearing, Social Distancing and the Predicted Probability of *R_t_* below 1 Projected values from a logistic regression model measuring the association of community transmission control (*R_t_* <1) with mask-wearing and social distancing in US states adjusting for population density, percent non-white and a time trend (Model 1). Values of social distancing are from the Google Community reports of relative residential time and represent no change from baseline, the national median during the study period (June 3 – July 31, 2020), and 97.5th percentile observed during the study period. Observed mask wearing was between 8.1%-73.7%, so estimates outside this range are model-based extrapolations. Horizontal line placed at 80% probability of community transmission control, though the desired % may be higher.

We evaluated the change in mask wearing in the 2 weeks before and after statewide mask mandates for 12 states (**Figure 4**). While there was a general trend of increased mask usage across these states over time, the linear segmented regression models resulted in no significant change in slope in crude, β = .04 [95% CI: -.47, .54], or weighted, β = .31 [95% CI: -.28, .91], mask usage following the interventions. There was a non-significant 2.2% [95% CI: -2.1%,6.5%] change in average mask usage following the mandate in the unweighted model and a 2.31% [95% CI: -2.76%,7.38%] change in the weighted model.

**Figure 4.**
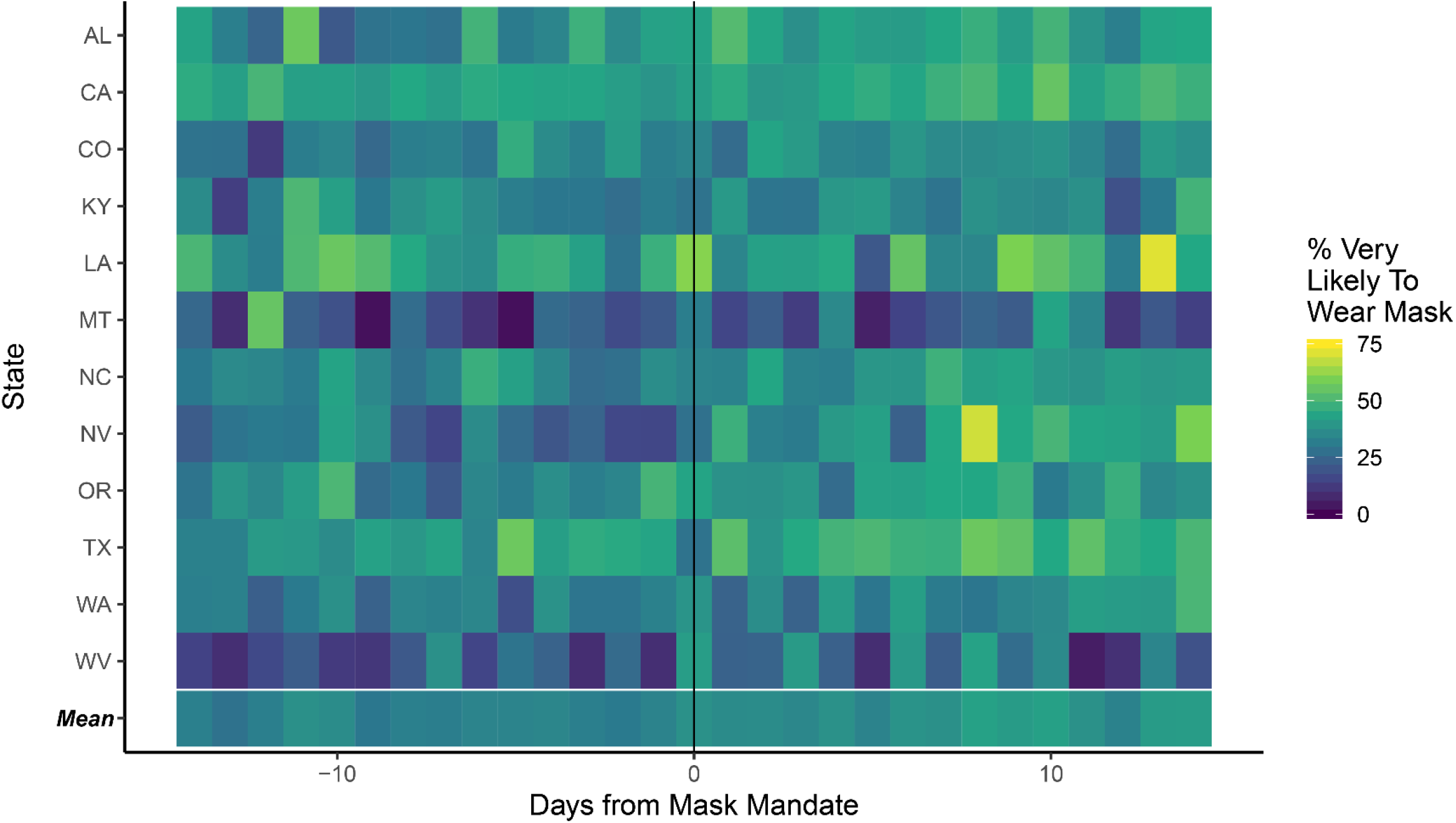
Mask Wearing in 14 Days Preceding and Following Statewide Mask Mandates Daily measure of the percentage of individuals that report they are very likely to wear a mask with family/friends and to the grocery store in 12 states. Values are for the 14 days preceding and following each state’s institution of a statewide mask mandate. The mean daily value across all states is presented for the same timeframe.

## Discussion

The effect of mask wearing on community transmission of SARS-CoV-2 has been the subject of substantial debate, despite evidence from detailed transmission studies and population-wide data from other respiratory pathogens. Here we present findings from over 300,000 serial cross-sectional surveys administered daily in June and July 2020 that confirm a high percentage of reported face mask wearing is associated with a higher probability of transmission control in U.S. states. Face mask wearing was more commonly reported among certain socio-economic groups (especially non-white and lower-income respondents). However, the impact of face mask wearing on lowering the instantaneous reproductive number below one – the threshold required for transmission control – remained despite adjusting for demographics, social distancing and prior peak transmissibility.

Mask wearing is shown to increase the odds of transmission control across all levels of social distancing, suggesting that any intervention to improve this community-based behavior may be worthwhile. The absence of a statistical change in mask wearing the two weeks following state-wide mandates highlights the point that regulation alone may not drive increased masking behavior. However, we found that there is a general increase in mask wearing prior to the implementation of these policies and mask mandates may be important tools in maintaining this trend. Masking behavior assessed from anonymous surveys may provide insight into where education or other interventions should be directed. These results are consistent with case studies of mask wearing in the United States^16,17^, and potentially provide some insight into why one report found no substantial effect of mask mandates in conjunction with other interventions^18^. The data presented here may highlight a gap between governmental policy and actual user behavior, but more research is needed.

Our evidence supports the role of mask wearing in controlling SARS-CoV-2 transmission; however, this ecological study cannot inform questions of causality. It is difficult to disentangle individuals’ engagement in mask wearing from their adoption of other preventative hygiene practices, and mask wearing may be serving as a proxy for other risk avoidance behaviors not queried (e.g. avoiding indoor spaces) – especially considering the magnitude of the reported effects. While our findings remained despite controlling for individual-level contacts from the Facebook survey, a proxy that likely includes other aspects of risk reduction, the possibility of residual confounding is likely. Additionally, observations from smaller states are overrepresented when results are aggregated at the state-level and further observations from survey respondents may not reflect the general population. However, our findings were consistent even with application of US census-based survey-weights.

A state that demonstrates social distancing may also be subject to additional non-modeled but impactful interventions including gathering size reductions, travel limitations, and the closing of businesses^38^. While the social distancing proxy used here captures the broad activity level that would result from the implementation of these policies, future research should focus on incorporating data from disaggregated interventions with empirical assessments of mask wearing. Additionally, potential *R_t_* and mask wearing within a state may be the result of prior transmission. While we showed the effect of mask wearing was robust to peak *R_t_* in the first wave of the epidemic, our methods do not control for time-dependent confounding or variations in mask usage by susceptibility status.

The validity of epidemiologic parameters of transmission are only as accurate as the incidence data to which the models are fit. If states reporting low mask wearing also underreport incidence (e.g. limited testing), we may be underestimating the true effect of mask wearing. Conversely, instantaneous *R_t_* estimations of transmission are subject to uncertainty, especially towards the end of the time-series before reports are complete^27^. While our results were robust to different estimators, our model does not account for estimation error. Additionally, mask wearing measures, social distancing and *R_t_* all exhibit significant temporal autocorrelation. To combat this, we dichotomized *R_t_* and aggregated our exposures by week, but further analyses may consider complex time-series models, mechanistic and quasi-experimental methods to estimate the effect of face masks.

We find a community benefit for “face masks” which collectively include masks of various hypothesized efficacies^39^. We did not query specific mask type or the use of face shields in conjunction with masks. The reported association may understate the maximum potential for face masks to curb respiratory transmission, which can only be realized through increasing the utilization of superior mask materials (i.e. choosing surgical masks over fleece gaiters). Additional study of effect modification by mask type is necessary to estimate true causal effects.

When considering the various challenges the US population has faced in slowing the spread of SARS-CoV-2, evidence on the impact of nonpharmaceutical interventions is paramount. Our data suggest widespread use of face masks by the general public may aid in limiting the SARS-CoV-2 epidemic as social distancing restrictions are rolled back around the United States. Given mixed evidence on the effect of mask mandates, but a strengthening body of evidence on the effect of masks, policy makers should consider innovative strategies for evaluating and increasing mask usage to help control the epidemic.

## Data Availability

Data will be made available

## Acknowledgements

The authors thank Kara Sewalk and Laura Wronski for their assistance. This work was supported by the Flu Lab. BMR and JSB acknowledge funding from Google.org and the Tides Foundation [TF2003-089662]. LFW acknowledges funding from the National Institutes of Health [NIH R01 GM122876]. CMA acknowledges funding from the National Institutes of Health [NIH K23-DK120899]. JS acknowledges funding from the National Science Foundation [DMS-2027369] and the Morris-Singer Foundation. MUGK acknowledges funding from the EU H2020 program MOOD and a Branco Weiss Fellowship.The funding bodies had no role in the decision to submit the manuscript for publication.

## Contributions

BMR, LFW and JSB conceived of the study. BMR, LFW, CMA and JSB wrote the first draft of the manuscript. BMR, LFW, MRB, and CMA contributed to the data analysis. JC, JB and JC oversaw and implemented data collection. All authors contributed to the interpretation of the data and editing of the final manuscript. All authors have seen and approved the manuscript.

## Supplementary Materials

**Figure S1.**
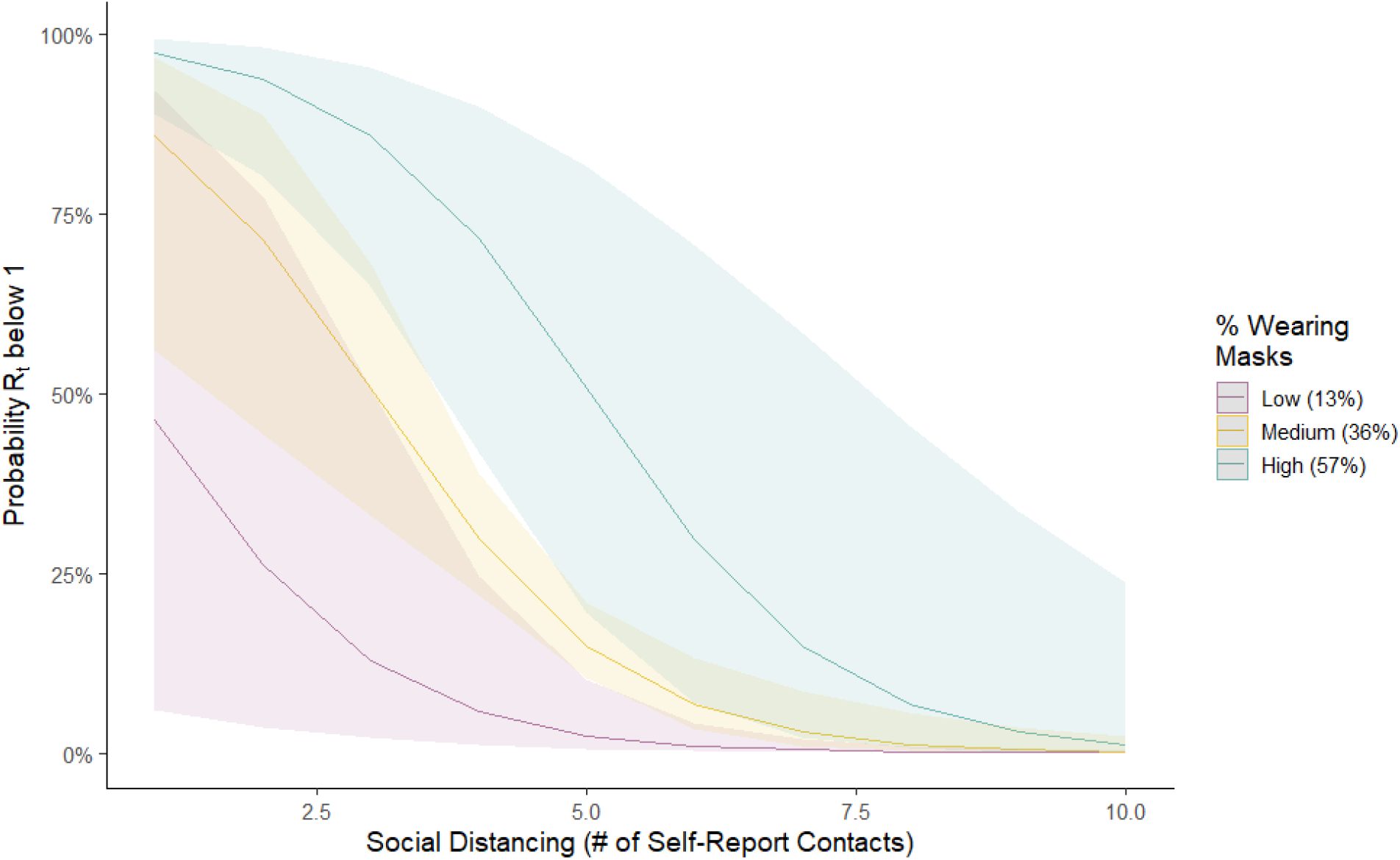
Mask Wearing, Social Contacts and the Predicted Probability of *Rt* below 1 Projected values from a logistic regression model measuring the association of community transmission control (*R_t_* <1) with mask-wearing and social contacts in US states adjusting for population density, percent non-white and a time trend (Model 1). The number of self-reported contacts at “social gatherings” from Facebooks’ COVID-19 symptom survey was aggregated over each week and state utilizing a weighted sampling scheme.

**Figure S2.**
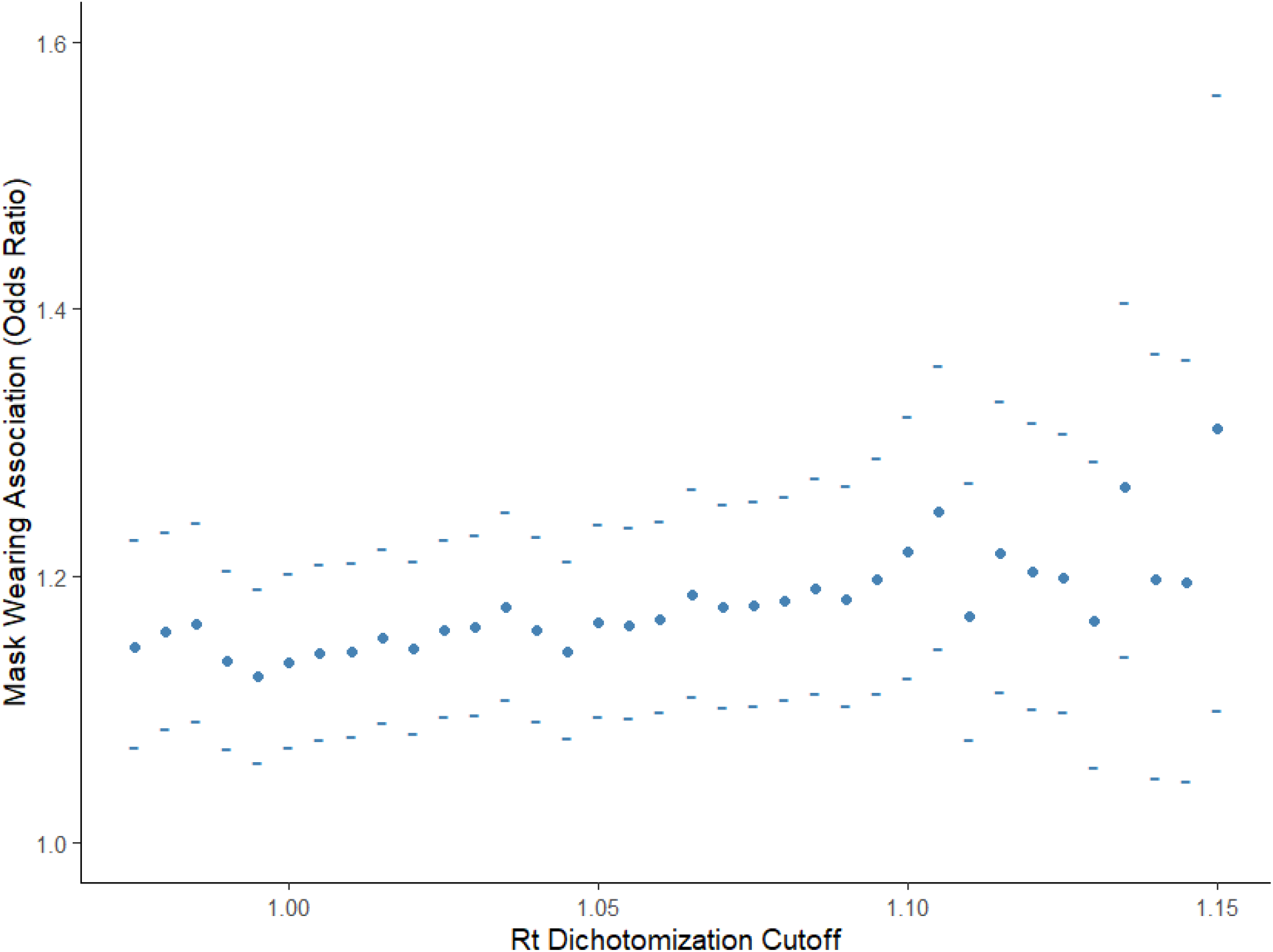
Association of mask wearing with *R_t_* at different dichotomization cutoffs Results from a logistic regression model measuring the association of community transmission control (*R_t_* < *x*) with mask-wearing adjusting for social distancing, population density, percent non-white and a time trend. Model was repeated as cutoff for *R_t_* dichotomization (*x*) was varied. The odds ratio (point) and 95% confidence interval (-) for mask wearing that resulted from each iteration is shown.

**Figure S3.**
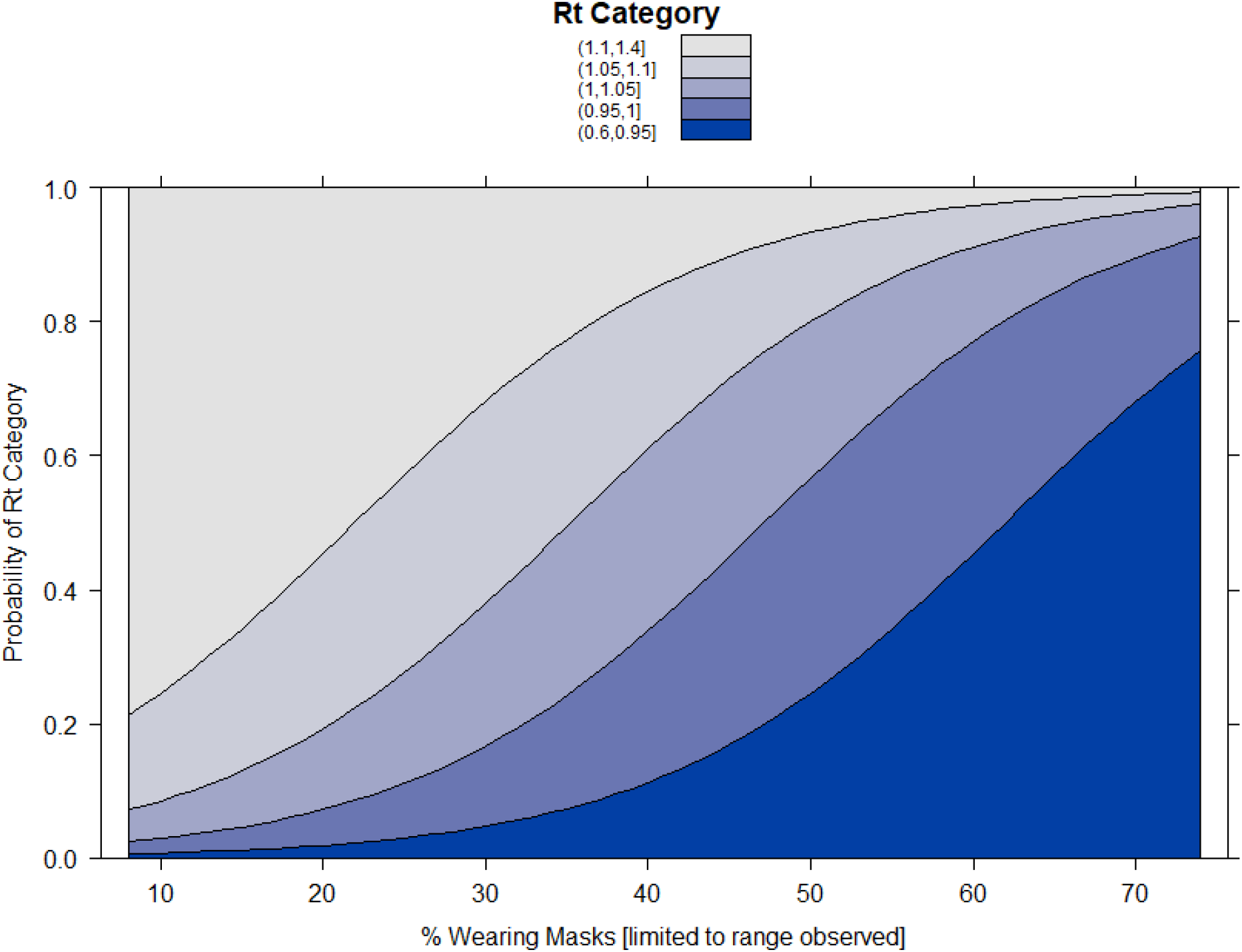
Association of mask wearing with categorical *R_t_* Projected probabilities from an ordinal logistic regression model measuring the association of community transmission control (*R_t_*) with mask-wearing adjusting for social distancing, population density, percent non-white and a time trend. Observed mask wearing was between 8.1%-73.7%.

